# Perceived Barriers and Enablers to Shared Decision-Making in Assessment and Management of Risk: A Qualitative Interview Study with Mental Health Professionals

**DOI:** 10.1101/2025.07.15.25331176

**Authors:** Nafiso Ahmed, Fareha Begum, Sally Barlow, Lisa Reynolds, Nicholas Drey, Kathleen Mulligan, Alan Simpson

## Abstract

**Background:** Involving service users in decisions about their care is essential to delivering recovery-orientated mental health services. Research shows, however, that service users often are not involved in the assessment and management of the risk component of their care. This paper explores mental health professionals’ perceptions of the barriers and enablers to shared decision-making in risk assessment and management.

**Methods:** We conducted semilJstructured interviews with fifteen mental health professionals working within either a community mental health or an early intervention service in the United Kingdom. Data collection and analysis were guided by the Theoretical Domains Framework for behaviour change, which encompasses fourteen domains identified as influencing behaviour.

**Results:** The findings show that staff are motivated to work collaboratively but experience a complex range of barriers that hinder the implementation of shared decision-making, including challenges related to the individual’s mental capacity and level of engagement, managing disagreements, type and level of risk, quality of therapeutic relationships, fear of causing distress or disengagement, and environmental constraints such as time, resources and place of meeting. Enabling factors included a supportive multi-disciplinary team, training they received as part of education and practice, clinical experience providing them with skills, and a belief that equipping individuals with knowledge and understanding of their risk and safety could help them to manage it more effectively.

**Conclusion:** The findings of this study may inform future intervention design to enhance collaboration in the assessment and management of risk and support current clinical practice in this area.

## Introduction

Mental health services are seeking to actively engage service users and their support network in decisions about their care and treatment (1–3). Shared Decision Making (SDM) is a collaborative process by which a healthcare professional (and carer if appropriate) support a service user to make decisions about their care and treatment (4, 5). SDM recognises service users’ unique expertise in their lived experience and fosters a sense of ownership in the recovery process (6). The implementation of SDM in mental health care is supported by policy (7), and widely valued by professionals and service users (8, 9).

Evidence of the effectiveness of SDM interventions for improving mental health outcomes is, however, inconclusive. A systematic review of SDM interventions for people living with mental health conditions found improvements in service user reported involvement in the decision-making process and minimal effects on consultation duration (10), though, the overall impacts on key outcomes—such as symptoms, readmission rates, participation, recovery, and satisfaction—were generally small with a low certainty of evidence. The authors of this study concluded that further research is needed to strengthen the evidence base in this area (10). Notably, most of the included studies in the review focused on decisions about medication and treatment. None addressed the use of SDM in the context of risk assessment or safety planning, suggesting that this remains an underexplored area in existing research.

There is ongoing debate in the literature about the feasibility of implementing SDM with individuals who are particularly vulnerable to paternalistic and coercive treatment, such as those who are acutely unwell or living with serious mental illness (SMI) such as schizophrenia, bipolar disorder, and major depression (11). A model of SDM in mental health, which conceptualises the process through three components—being informed, involved, and influential—highlights that implementation can vary depending on factors such as the clinical setting, mental capacity, and the individual’s desire to participate in decision making (12). While the model recognises that participation may vary, studies have shown that people with SMI often desire to be involved in decisions about their care and treatment (13, 14) and can actively participate when supported appropriately (15, 16).

Another contentious area for SDM is where there are issues around risk and safety. Risk and safety considerations are central to decision-making in mental health services, and Mental Health Professionals (MHPs) are key in mitigating potential harm to self and others including suicide, self-harm, and violence (17, 18). Risk assessment is the mechanism by which MHPs identify safety concerns, which usually involves gathering information through either clinical judgment, structured assessment tools, or a combination of both, known as structured clinical assessment (19).

The National Institute of Clinical Excellence (NICE) guidance explicitly states that risk tools and scales should not be used to predict suicide or self-harm, or to guide treatment or discharge decisions (2). This recommendation is echoed in recent guidance for suicide prevention (20) and assertive and intensive community mental health care (21). Instead, these guidelines advocate for relational, person-centred approaches to safety (22), as risk prediction tools, scales, and stratification have been found to lack predictive accuracy (23).

A risk management plan includes strategies for effectively reducing or managing the identified risks from occurring (17). The implementation of these plans in practice can be complex, particularly in balancing risk while respecting a service user’s autonomy (24). Research suggests that MHPs often perceive risk management and recovery orientated care as competing priorities (25). Nonetheless, best practice guidelines emphasise the importance of collaborative risk management practices, involving the professionals, service users, carers (where appropriate), and the wider multidisciplinary team (2, 17, 20).

Doyle, Grundy (26) have coproduced a set of updated best practice principles in clinical risk management, reinforcing the need for person-centred approaches where service user involvement is central. Their paper supports the shift from risk-averse practices towards partnership-based strategies that balance safety with individual empowerment. They highlight the importance in providing professionals with support through regular supervision, and the opportunity to learn from incidents without blame.

Although shared decision-making is a cornerstone of person-centred care, its implementation in the assessment and management of risk for individuals with SMI remains inconsistent (27). Many service users have reported being unaware of the content and purpose of their risk assessments (25), or a lack of involvement in identifying and managing risk (28), thus, limiting their ability to engage meaningfully in decision-making. Barriers to SDM identified in our previous interview study with service users include lack of awareness of the risk assessment and management process, power imbalances between professional and service user, and difficulty in discussing sensitive topics.

Perceived enabling factors identified included the opportunity to contribute to the identification and management of risk, to enhance the ability to maintain their own well-being and safety and to gain a better understanding of risk issues (29).

In our previous systematic review (27), we sought to identify literature on MHPs’ perceived barriers and enablers to SDM in risk assessment and management. While all studies included in our review addressed at least one component of the’Three Is of SDM’ model—Informed, Involved, and Influential (12) — none of the studies examined SDM directly. A significant gap in the literature therefore remains regarding our understanding of the implementation of SDM in the assessment and management of risk.

This current study aims to fill this knowledge gap by identifying specific factors that MHPs perceive as either hindering or facilitating SDM in this context. The findings may contribute to improvements in practice by highlighting areas or elements of SDM that may present challenges, and to support future research efforts to enhance collaboration with service users in the risk assessment and management process. This study aimed to address the following research question:

*What do mental health professionals perceive to be the barriers and enablers to shared decision-making in the assessment and management of risk with people with severe mental illness?*

## Methods

This study utilised a qualitative research design to interview mental health professionals about their experience of SDM in the assessment and management of risk with people living with SMI. A detailed description of the study methods can be read in our earlier publication on service users’ experiences (29). The of this study adheres to the Standards for Reporting Qualitative Research (SRQR) 21-item checklist (30).

### Participants

Mental health professionals were recruited from two Community Mental Health Teams (CMHT) and an Early Intervention Service (EIS) in one NHS trust serving an inner-city area in England. Potential participants could take part in the study if they were senior practitioners (e.g., managers or psychiatrists) or care-coordinators (e.g., social workers, occupational therapists, or mental health nurses) working within Community Mental Health Teams (CMHTs) or Early Intervention Services (EIS) and were responsible for conducting risk assessments and risk management A stratified purposive sampling strategy was used to identify and invite mental health professionals for interviews. The team administrator for each setting provided a staff list that included professional background and role. Five MHPs representing senior practitioners and care-coordinators were selected from each staff list and emailed an invitation to interview. Reminder emails were sent after a week. If the professional failed to respond to the reminder email or declined the invitation to participate, the next person from the same professional background was selected and emailed an invitation to interview.

### Material

The interview schedule was developed using the Theoretical Domains Framework (TDF), a comprehensive framework designed to identify and understand factors influencing behaviour change (31, 32). The TDF integrates 33 psychological constructs from multiple theories into a single validated framework (33) comprising 14 theoretical domains. Table 1 presents a list of all 14 domains, along with their definitions.

**Table 1.** TDF domains associated belief statements and illustrative quotes.

Questions were developed in collaboration with a TDF expert (KM), and previous studies that had applied the TDF were referred to (34–36). The interview guide was piloted with a clinical academic experienced in conducting risk assessment and management with individuals with SMI. The interview schedule started by asking participants about the meaning of risk and SDM, followed by the processes, frequency, and challenges associated with assessing and managing risk in severe mental illness (Additional file 1). The current paper reports on the findings that relate to potential barriers and enablers to SDM in these practices.

### Data collection

Semi-structured interviews were carried out from January to October 2017 and lasted between 22 and 60 minutes. All interviews were conducted by NA in a private office space at the CMHT/EIS site.

Interviews were recorded using a digital audio device and were transcribed verbatim professionally. The transcriber of the recordings signed a confidentiality agreement, and NA checked the transcripts against original recordings for accuracy and any identifying details were removed. Demographic information was collected from each participant using a brief questionnaire adapted from a previous study (37).

### Data analysis

Anonymised transcripts were imported and managed using QSR International’s NVivo 11 qualitative data analysis software (38). Analysis drew on established methods used in previous literature (34–36, 39), employing a deductive thematic approach guided by the TDF (31) and involved the following six stages:

#### Step 1: Developing a coding guide

A coding guideline was developed based on the 14 domains and 84 constructs of the Theoretical Domains Framework (33).

#### Step 2: Pilot coding exercise

Two authors (NA and FB) jointly coded findings from two randomly selected interview transcripts, and the coding guide was refined and finalised.

#### Step 3: Coding and assessing reliability

All the remaining interview transcripts were then independently coded by the same two researchers (NA and FB) into the 14 TDF domains using the coding guide. Coder reliability was assessed for the last four transcripts using percentage/agreement (35, 36, 40, 41).

#### Step 4: Thematic synthesis and generating specific beliefs

Belief statement representing the specific underlying belief for each participant’s response within each theoretical domain were generated. A belief statement is defined as *‘a collection of responses with a similar underlying belief that suggest a problem and/or influence of the beliefs on the target implementation problem’* (31). Therefore, responses with similar underlying themes were grouped, and a summary belief statement was generated. New belief statements were created for responses that could not be grouped. A frequency count for each belief, capturing the number of participants who mentioned the specific belief in their response, was calculated.

#### Step 5: Identifying relevant theoretical domains

In line with previous publications (34, 35), the TDF domains relevant to the target behaviours were identified. Domains were identified as relevant if they contained a specific belief (step 4 above) that might be a potential barrier or enabler. In addition, three factors were considered when identifying the key domains: frequency of belief across interviews, presence of conflicting beliefs, and evidence of strong beliefs that might impact the behaviour. All these factors were considered simultaneously to establish the relevance of the domain in influencing the target behaviour.

#### Step 6: Validating the mapping of beliefs statement to domain

To ensure belief statements accurately represented the domain, a researcher with experience using the TDF (KM) validated each belief statement. Blinded to the domain in which the belief statement had been generated, KM was asked to assign each belief statement to a TDF domain. The agreement between the two researchers (KM and NA) was calculated through the number of items on which the two coders agreed divided by the total number of items, multiplied by 100.

#### Ethics

Ethical approval was granted by the NHS Health Research Authority Research Ethics Committee London—Camden & Kings Cross (16/LO/1918). A substantial amendment was submitted to gain approval to increase the sample size, include a second CMHT site, to advertise on the notice boards in the CMHT/EIS centre. The same research ethics committee granted a favourable ethical opinion of the amendment.

## RESULTS

### Response rate and participants’ characteristics

Twenty mental health professionals were invited to take part, and a total of fifteen agreed to participate (75%). The sample consisted of senior practitioners (n=4, 27%) and care coordinators (n=11, 73%) from various professions. The majority were nurses (n=7, 47%) and social workers (n=4, 27%) and had been working within their current community mental health service for over ten years (n=6, 40%). Most professionals worked within a CMHT setting (n=10, 67%) compared to EIS (n=5, 33%) and were female (n= 12, 80%). Demographic information is presented in Table 2.

**Table 2.** Mental health professional participants’ characteristics.

### Findings

All the TDF domains were mentioned by at least one participant. Knowledge, Skills, Social Professional Role and Identity, Beliefs about Capabilities, Optimism, Reinforcement, Goals, and Social Influences were mentioned by all 15 participants (Additional file 2). All domains were supported by the interviews, the most frequently identified were Social Influences and Reinforcement. Behavioural regulation was mentioned by the fewest number of participants and supported by the least number of quotes.

### Belief statements

A total of 75 belief statements were created across the 14 domains, ranging between 1 and 17 per domain (mean = 5.4, SD = 4.2). See **Error! Reference source not found.** file 2 for full details of all 75 belief statements and the frequency with which they were coded. After discussion within the research team 48 belief statements across all 14 domains, between 1 and 7 per domain, were deemed most relevant based on the criteria described in step 5 of analysis. For example, within the ‘social influence’ domain, the belief statement *‘The service users’ capacity is a key factor in whether I can implement SDM in risk assessment and management’* was deemed relevant as it was frequently mentioned both across and within interviews. Table 1 details the relevant beliefs, with example anonymised quotations taken directly from the transcripts.

### Inter-rater reliability

Based on previous research (36), inter-rater agreement was calculated for the last four transcripts (27%). Across the 14 TDF domains, inter-rater agreement between the two coders ranged from 84 % to 100 %. When blinded, a TDF expert was asked to map the relevant belief statements onto a domain. For 34 (71%) of the 48 beliefs the expert mapped the belief statement onto the intended domain, whereas for 14 (29%) beliefs the expert mapped the belief statement onto a different domain. Two researchers (NA and KM) met to discuss and reach a consensus. After discussion, they agreed that 41 (85%) beliefs should remain mapped to the intended domain and seven (15%) beliefs were remapped to the domain identified by the expert.

### Beliefs mapped to the TDF domains

#### Knowledge

Over half of the participants (n=8, 53%) referred to guidance on the implementation of shared decision making in risk assessment and management, including from their local trust policies, the Care Programme Approach (CPA), the Care Act (2014), the Mental Capacity Act (2005), and/or the National Institute for Health and Social Care Excellence (NICE) in England and Wales. In contrast, seven participants reported being unaware of any guidelines or policies specifically recommending SDM in the risk assessment and management process.

#### Skills

Although most of the professionals (n=10, 67%) interviewed acknowledged that discussing risk could be difficult, many noted that strong communication skills aided them in addressing sensitive topics related to risk with both the service users and others involved in the SDM process. Several professionals reported adapting their language to facilitate discussion about risk, for example, by rephrasing questions, or judging how best to communicate risk information based on the service user’s understanding and willingness to engage.

A majority of professionals (n=11, 73%) believed that confidence in discussing risk with service users increases with clinical experience. One professional highlighted that ‘old fashioned views’ that were less enthusiastic about service user involvement could impede SDM, whereas another suggested that the experience required to implement SDM depended on context. For example, in decision-making about daily living, such as using a cooker, an Occupational Therapist (OT) student might have more relevant experience to conduct the assessment with the service user than a psychiatrist.

About half of the participants (n=7, 47%) believed that a qualified and trained professional was required to implement SDM in risk assessment and management with service users. Some referred to the training they received in their professional education enabling them to support service users in managing their risks. Others highlighted local training or Approved Mental Health Professional (AMHP) training preparing them for collaborative risk assessment and management. A senior professional spoke about newly qualified professionals being trained to talk more openly about risk with service users and compared this to the paternalistic training that they had received.

Two thirds of professionals (n=10, 67%) expressed the belief that additional training could enhance the implementation of SDM in risk assessment and management. Training needs included knowledge about risk assessment and management frameworks; policies and procedures on how best to implement SDM; and exploring the different meanings of risk. Professionals believed that for training to be effective, it needed to be frequent, attended together as a team and co-facilitated by service users or other services such as the police.

Professionals valued gaining their colleagues’ perspectives, support, and reassurance about how best to assess and manage risk with service users. They said forums were available to them where they could talk openly and share concerns with colleagues, including case discussion groups or ‘morning traffic light meetings’. They described the traffic light system as a daily open forum between the Multi-Disciplinary Team (MDT) where each service user risk level was categorised as low, medium, or high. The service users deemed high risk were discussed with the MDT.

Therapeutic, interpersonal, decision-making, and listening skills were also identified as essential in facilitating shared decisions about risk with service users. Professionals also highlighted key personal qualities including patience, empathy, a non-judgemental attitude, compassion, and honesty.

#### Social/professional role and identity

A majority of professionals (n=11, 73%) perceived risk assessment and management as a collaborative process, emphasising shared responsibility between themselves and the service users to work together in minimising risk. They regarded their role as empowering service users to take part in the RA and RM process, informing service users about their risk, and teaching them skills to protect themselves. Many talked about risk documents belonging to the service user and decision-making being “about them”, therefore, they described their role as supporting service users to take the lead or to contribute to the process. Professionals emphasised the need to move away from the’medical model’ or the ideology that they are the’expert’ who are solely responsible for treatment. Instead, they advocated for a more collaborative approach where the service user has a role in taking ownership for and engaging in identifying and reducing risk.

Most participants (n=10, 67%) acknowledged the lead professional (i.e., care-coordinator) as responsible for facilitating SDM in the process of assessing and manging risk, highlighting that the care-coordinator possesses the skills, training, and knowledge to liaise with the multiple parties involved, to coordinate discussion and manage anxieties about risk. Many (n=8, 53%) also considered risk assessment and management as a team responsibility where everyone involved in the individual’s care had a role in implementing SDM.

Although professionals perceived implementing SDM in the risk assessment and management process as important, they said their duty of care to protect the individual and/or others from risk took priority. A few professionals (n=7, 47%) described situations where they made decisions on behalf of the service user or when certain risk information was not shared. These actions were typically framed as being in the person’s best interest, particularly where the individual was assessed as lacking the capacity to understand or refused to engage. Professionals referred to the legal framework of the Mental Capacity Act (2005) or their statutory obligation to safeguard the individual. For example, a senior professional explained that when a service user refused to accept risk management advice, as the responsible clinician, they sometimes needed to act on the service user’s behalf to protect them or others from risk. The professional described this as challenging and as a last resort, as they felt it could be debilitating and patronising for the service user.

Some (n=6, 40%) felt that SDM was already embedded in their practice and that of their team, others, however, believed that there was scope for improvement and situations where service users were not being involved.

#### Beliefs about capabilities

Most professionals (n=11, 73%) said they felt confident in implementing SDM in risk assessment and management with service users. Confidence was increased by training, experience, team structure and receiving support from colleagues (further detail in skills and social influence domain). One professional reported a lack of confidence in discussing “risk to others” with service users, and a few highlighted that their confidence depended on factors such as the service user’s reaction to the risk discussion, current presentation and their ability to reach a shared decision.

#### Optimism

Most professionals (n=12, 80%) felt optimistic that they could implement SDM in risk assessment and management in the future, some reported that it was already happening and thus felt positive it would continue. Others expressed uncertainty about being able to implement SDM in risk assessment and management in the future. For example, a participant stated that their workload was becoming increasingly busy and stressful, and that SDM takes time.

#### Beliefs about consequences

Professionals acknowledged that risk was an emotive topic for some service users and that discussing distressing periods of their lives was understandably difficult. As a result, professionals questioned the helpfulness in having conversations about risk history, and others highlighted the need to consider the potential consequences for the service user by having these conversations, such as causing distress, or setting them back.

Often the behaviours and risk issues that needed to be discussed as part of the risk assessment process were believed to have occurred when the service user had limited awareness or insight. Professionals were, therefore, concerned that discussing risk history as part of the risk assessment process or sharing these documents could trigger a reaction, such as alarm, embarrassment, or resistance to accept that this is how they presented.

Many professionals (n=9, 60%) worried that a service user could become offended by a discussion about risk and that this could cause them to disengage from services. For example, a professional described how a service user disengaged from services when they realised that the professional knew about their risk history (e.g. criminal conviction) and were worried that their family would find out.

Professionals acknowledged societal misconceptions associating mental illness with dangerousness and thus were conscious about compounding stigmatising beliefs by focusing on risks, recognising that a service user’s historical risks could remain on their record for years and cause them to experience labelling and stigma. For example, a professional said that a service user might be restricted from accessing certain services because of their risk history, therefore, they emphasised the need to update the risk assessment regularly with the service user.

Nonetheless, several professionals (n=6, 40%) considered implementing SDM as an opportunity to build service users’ awareness of risk and to support them in managing their risks. A professional explained that an individual, for example, who had been abused their entire life may not perceive themself as at risk and therefore highlighted the importance in discussing risk with individuals to help them with identifying and minimising that risk. Another professional talked about a service user who smoked heavily, not understanding the health consequence of smoking, the professional felt it was their responsibility to provide the service user with knowledge of the risks of smoking and to discuss potential interventions with them.

Professionals believed that collaborating with a service user in risk assessment and management could reduce the likelihood of the risk occurring. When a service user was involved in the process, professionals felt that they were more likely to put a plan in place that the service user understood and engaged with.

#### Reinforcement

All the professionals interviewed in this study endorsed SDM in risk assessment and management with service users. Nearly all (n=14, 93%) claimed that recovery, person-centred care or promoting empowerment encouraged them to implement SDM in these practices. Professionals highlighted that collaborating in these processes provided the service user with an opportunity to share their own experiences with risk, as they are experts about their own lives and know the most about their risk history. A few professionals believed that the absence of service user involvement in risk assessment or management would confine the process to a “paper exercise” making it “meaningless”.

Furthermore, service users being involved and owning the risk decision was believed to increase adherence to the risk management plan.

Being honest and keeping the service user informed about the content of the risk assessment motivated professionals to implement SDM. By being transparent about risks and sharing the document, professionals felt that a service user’s self-stigma and anxiety towards their risk history could be reduced. Professionals recognised that some service users were not aware of the risk information held about them and said that if it were them, they would want to know what was recorded on their record. Professionals who worked within EIS said that engaging in open dialogue was the way they formed relationships with the service user. They believed that being open and honest about risk could prompt service users to disclose risk issues that were concerning them.

Safeguarding and protecting the service users and others from risk encouraged many professionals to implement SDM. Professionals talked about being trained in helping service users to manage their risks and protect themselves, and this motivated them to implement SDM. For example, a professional spoke about making a service user aware of the possibility of accidentally harming someone (or being arrested) by carrying a knife for protection. If the service user did not accept the information provided or lacked capacity to understand, the professional spoke about the need to manage that risk without their collaboration.

#### Intention

Two thirds of professionals (n=10, 67%) felt that SDM in RA and RM was already embedded in their practice and that of their team and expressed positive attitudes about this continuing. Others supported SDM in theory but recognised that it was not always happening in practice. They stated that this was something that their team were working towards. A senior professional acknowledged that risk assessment often focuses on risk to self and others and said risks such as stigmatisation and isolation could be considered more with the service user.

#### Goals

All professionals believed it was important to implement SDM. However, the majority (n=10, 67%) emphasised the value of providing service users with knowledge and understanding about their risks, as to increase both awareness and ability to manage risk.

Additionally, over half said that they were motivated to discuss risk with a service user for information gathering purposes. They believed exploring risk directly with the service user was key in the initial assessment process for newly referred service users, who often had an unknown risk history. Participants talked about the formal assessment process where they would gather information from the service user by asking a series of questions about their understanding and experiences with risks. Even with existing service users, professionals stressed the importance of verifying the recorded risk information by talking with the service user. Professionals believed engaging in an open dialogue about risk could encourage the service user to volunteer unknown risk information.

Achieving a shared decision about risk and how to manage risk with the service user was valued by professionals (n=6, 40%). They believed that achieving SDM could potentially reduce the risk from occurring. Collaborating with the service users in the process would also mean that they would share the responsibility for risk, and also the service user’s perspective of risk would be included.

Professionals felt that this was the right move towards personalised care.

Other factors relating to’goals’ that motivated professionals to implement SDM in RA and RM included developing an advanced directive with the service user and improving the therapeutic relationship (discussed within the “social influences” domain).

#### Memory, attention and decision processes

The type of risk influenced professionals’’decision about whether or not to implement SDM in risk assessment and management. Professionals perceived “risk to others”, such as violence and aggression, as more difficult to discuss with service users compared to “positive risk” and “risk to self”. When the risk was imminent and potentially harmful, professionals said that they may not have the chance to involve the service user in the decision-making process. Professionals said that they were more likely to involve service users deemed as high risk in the RA and RM process compared to service users with relatively few risks.

Professionals identified individual factors relating directly to the service user as barriers to implementation. For each individual, they judged whether it was appropriate to discuss risk and the consequences of discussing risk, therefore, they considered a blanket approach to implementing SDM as inappropriate, as they believed it depended on the context of the risk and the individual.

#### Environmental context and resources

The format of the risk assessment document enabled some professionals to implement SDM. For example, professionals spoke about a new care planning document including a safety section that needed to be completed with the service user.

On the other hand, several professionals felt that they did not have adequate time to implement SDM due to increasing workload. They suggested that lower caseloads and time allotted at care planning meetings could support them to implement SDM in risk assessment and management. Professionals spoke about constantly managing crises and trying to keep the service user and others safe, which they believed limited their time to implement SDM properly. Others highlighted a lack of resources and the opportunity to get all parties in a room to discuss risk as barriers to SDM. A senior manager, however, argued that a lack of time or resources was not always a good enough reason to not involve service users in the decision-making process, as the service user could be contacted via telephone instead.

The team structure and setting type influenced professionals’ ability to implement SDM in risk assessment and management with service users. A professional working within one of the CMHTs explained that decision-making about risk in their setting was often discussed in a team environment. The service user presence, however, in the decision-making process was absent from the professionals’ response indicating that SDM to them may signify working collaboratively with other members of the MDT. Another CMHT professional compared their current service to forensic mental health services and suggested that forensic services had clearer guidelines about implementing SDM in risk assessment and management practices.

Professionals working within early intervention service (EIS) accredited their ability to implement SDM in risk assessment and management to their setting type and team dynamic. They identified their traffic light system (described earlier in ‘Skills’ domain) as helping them to work collaboratively with service users in managing their risk, as responsibility for risk was shared amongst the team and risk regularly discussed. As a result, professionals felt supported in implementing SDM with service users.

Professionals working within the EIS also spoke about engaging in family therapy with the service user, carer and psychologist, and this being an opportunity to discuss risk in a supportive environment. Professionals recognised that their EIS structure was developed on the recovery model, so felt that their practice embedded the ethos of empowering service users to manage their own mental health and safety. Other aspects of the EIS that enabled professionals to implement SDM in RA and RM were their specific relapse prevention plans, smaller caseloads, and EIS being the first point of contact for service users. One participant in EIS, notably, attributed their ability to implement SDM to the openness within their team in assessing and managing risk and the reduced hierarchal structure that allowed them to feel respected, and their opinions validated.

The location where the RA meeting took place influenced professionals’ ability to discuss risk with the service user (n=6, 40%). Professionals found it easier to address risk with service users at their site offices, which had panic alarms in case the service user did not respond well to the conversation about risk. Some professionals said they were reluctant to address risk openly with a service user in their home environment, especially when they did not know enough about the service user’s risk history; they had concerns about the service user’s presentation or home environment and feared for their own safety. Other professionals identified their buddy system, working in pairs and carrying their sky guard (alarm system) as enablers to addressing risk with a service user in their home environment.

#### Social influences

All the interviewed professionals identified the service user lacking in capacity as a barrier to implementing SDM in RA and RM. Some reported a lack of capacity as the only factor that would prevent them from involving the service user in the risk assessment and management process. While some referred to the Mental Health Act being used to make best interest decisions, others talked about consulting with carers or advocates to support when the service user was lacking in capacity. A few professionals acknowledged the service user’s freedom of choice to make’unwise’ decisions and best interest decisions only being made when the service user was deemed as lacking in capacity.

Service users’ level of insight or awareness was identified by most professionals (n=14, 93%) as a barrier to SDM in risk assessment and management. Professionals talked about the service user not accepting the risk issue, or disputing the risk information recorded on their risk assessment, which impeded achieving a joint agreement of the problem and a shared understanding of how to manage it. Professionals talked about the difficulty in discussing risks that occurred when the service user was acutely unwell, as some individuals may have limited memory of what happened. Therefore, professionals recognised that going through historical risks with them could cause alarm or a reluctance to accept that this was’about them’, or emphasised timing and deciding when best to approach discussions about risk with service users. How the service users presented also determined whether it was appropriate to discuss risk. For example, a professional explained that if the service user presented as chaotic, aggressive or under the influence of alcohol or drugs, they were less likely to share risk information or to start a discussion about risk.

The willingness of the service user to take part in the risk assessment and management process was also perceived by professionals as a barrier to SDM. Professionals reported that some service users were disinterested in taking part in decision-making, or reluctant to engage in discussions about their risk history. Therefore, they sometimes needed to continue without the service user’s involvement and put plans in place to manage risk.

Professionals considered potential disagreements between themselves and the service user about risk as a barrier to SDM in risk assessment. They reported challenges in resolving disagreements with service users who refused to accept their recorded risk history, or became defensive when asked questions about risk. Consequently, professionals were worried about damaging the therapeutic relationship, compounding stigma, and causing distress. Other participants, however, emphasised the service user’s freedom of choice to disagree with their viewpoint and the importance in collaborating to reach a compromise. Professionals stated that the risk management plan was more likely to be effective if developed collaboratively and with the agreement of the service user. They considered engaging in an open dialogue about risk with service users as important despite disagreements.

Across services, professionals talked about multidisciplinary working and adopting a team approach as enabling SDM in the process of assessing and managing risk with service users. Meetings about risk with a service user usually involved a few members of the MDT, the service user and the carer, all contributing to the decision-making process. Professionals said they valued their colleagues’ advice, perspectives, and expertise in the decision-making process. For example, some considered it imperative to gain other professionals’ views on the final decision before they implemented it. Also, they considered it helpful to have another professional in the room that could potentially explain risk information more effectively to the service user.

A team approach to managing risk facilitated information sharing between the MDT, service user and carer. They considered this vital as when the lead professional was absent, there were other members of the team able to step in that were familiar with the case and the agreed plan. Professionals talked about the decision not only being theirs, but a decision that was shared with other members of the team. They felt that a team approach to assessing and managing risk would mean that the service user did not feel like only one professional was “harping” on about their risk, and that if something went wrong, accountability would be shared.

Professionals considered carers key in implementing SDM with service users. Participants mentioned contacting carers, with the service user’s permission, when the individual was experiencing a crisis or for information gathering purposes. There were, however, some accounts of carers impeding SDM in risk assessment and management with service users. For example, when the carer was believed to pose a risk to the service user, professionals highlighted the difficulty in reaching a shared understanding and managing the risk as the service user did not want to get their carer’in trouble’. Also, professionals talked about carers disclosing risk information about the service user and not wanting the professional to reveal where they had received this information. This caused difficulty for the professional in assessing and managing risk with the service users, as they had to consider the carer’s safety.

Professionals believed that their ability to implement SDM in risk assessment and management with service users depended on the quality of the therapeutic relationship. A close therapeutic relationship enabled some professionals to broach risk with service users. Others, however, talked about the difficulty in discussing risk with a service user who they did not have a rapport with, e.g., a newly referred service user. They explained the need to build trust and a therapeutic relationship before discussing risk. Conversely, damaging the therapeutic relationship by talking about risk with some service users caused concern for some staff.

Interprofessional conflict and power dynamics within the team acted as a barrier to shared decision making. For example, a CMHT professional cautiously reported hierarchal and power dynamics within their team, which they believed resulted in care coordinators feeling disempowered and discouraged from suggesting an appropriate risk management strategy for the service user. In contrast, an EIS professional complimented their shared team approach to assessing and managing risk. Their team’s reduced hierarchal structure aided them in decision making, as their consultant respected their opinion as the care coordinator.

A couple of professionals reported language as a barrier to implementing SDM in risk assessment and management, as SDM requires all parties to understand the information available. However, where there is a language barrier implementing SDM can be difficult, although one professional recognised that an interpreter could be used to overcome this.

#### Emotions

Professionals said that discussing risk with service users could sometimes cause them to feel anxious, particularly when addressing risks such as “exploitation” or “harm to others”. The possibility of a negative outcome, such as damaging the therapeutic relationship, contributed to professionals’ apprehension. Some noted that those with less experience in having these conversations with service users may feel especially nervous. Additionally, professionals expressed fear that a service user could harm themselves or others, which heightened anxiety around discharge planning or managing those risks. A few professionals described discussing risk with service users (and carers) as uncomfortable.

Some professionals (n=6, 40%) expressed fear for their personal safety in initiating discussions about risk or sharing risk information with some service users. During home visits, professionals were reluctant to discuss sensitive risk topics or to share risk information if they were concerned about the service user’s capacity, presentation, behaviour, or if it was the first meeting with the service user.

Other professionals were concerned about the service user becoming offended at the decision being made and potentially attacking or threatening them. Some differentiated between the inpatient and community settings in how they managed potential risks towards staff. They described ward staff having access to a response team to support them if the service user became angry or violent about the risk in discussion.

A manager also described the need to consider the person who was addressing risk with the service user, as some service users may have made threats in the past towards certain staff members, e.g., the consultant. They talked about being open with a service user about risk but ensuring that the content of the discussion was not putting another person at risk. The manager concluded that it was important to judge how and when to have conversations about risk and whether to seek support from another colleague.

Although professionals acknowledged the negative emotions they experienced when assessing risk with service users, many spoke about overriding these emotions as they believed discussing risk with the service user and involving them in the process was imperative. They discussed the need to manage their own emotions and to avoid becoming overwhelmed by others’ emotions.

A few professionals reported experiencing positive emotions from implementing SDM with service users, feeling hopeful and satisfied that they were doing the right thing by supporting service users to take charge of their own care.

#### Behavioural regulation

One participant suggested that a mobile app where the individual could access and contribute to their risk assessment could support shared decision making.

## Discussion

The main aim of this study was to explore what mental health professionals perceive as the barriers and enablers to shared decision-making in the assessment and management of risk with people living with severe mental illness. Key barriers identified through the qualitative interviews included the individual’s mental capacity and engagement, challenges in managing disagreements about risk, the type of risk being discussed, the quality of the therapeutic relationship, and fear of negative consequences such as causing distress, disengagement, or reinforcing stigma. Conversely, enabling factors included a supportive multidisciplinary team, training received as part of education and practice, clinical experience, a commitment to promoting empowerment and person-centred care, and efforts to enhance service users’ understanding of their own risk and safety to help them manage it more effectively.

While professionals expressed positive attitudes towards involving service users in the assessment and management of risk, they also conveyed reservations – primarily due to service user-related factors. A key barrier identified was a lack of mental capacity, with some professionals highlighting this as the sole reason they would refrain from implementing SDM in risk discussions. This finding aligns with our published systematic review (27) and the broader literature on SDM in mental health care (8, 42).

Stacey and colleagues model of SDM recognises that supporting service users’ influence in decision-making is the most challenging for mental health professionals (12), often due to concerns about mental capacity. The authors argue that even when it is decided that a best interest decision needs to be made, the service user should remain informed and involved in the process wherever possible (12). Service user’s influence in decision making can also be supported using decision aids and advance directives/choice documents, including with people with SMI (15, 43). A previous study (44) that evaluated the feasibility and effects of a SDM intervention for in-patients with schizophrenia found that it was feasible for most patients, and did not require more time. There were also positive effects found including increased perceived involvement in medical decisions, and uptake of psychoeducation.

Some professionals in this study felt that their ability to implement SDM in risk assessment and management depended on the quality of the therapeutic relationship, while others feared that discussing risk with service users could undermine it. These findings support previous research from the service users’ perspective, which highlights the importance of trust and a strong therapeutic relationship in fostering openness during risk management (45). The therapeutic alliance between mental health professionals and service users is important in delivering high-quality care and consistently linked to positive outcomes across a wide range of interventions and settings (22, 46, 47). However, risk management practices may negatively impact recovery-orientated care (48), therapeutic relationships (49) and risk assessment (50), underscoring the need for strategies that balance collaborative decision-making with effective risk management.

Half of the mental health professionals interviewed in the current study said they did not know of any specific guidelines or policies that recommended SDM in risk assessment and management. The other half recalled guidance in local and national policies but were largely unable to elaborate on its content. As a result, the majority (10/15) expressed a need for additional training in how best to implement SDM in their practice. A lack of training has been reported as a barrier to discussing suicidality with service users (51, 52) and engaging service users in risk management (53).

Educational initiatives where professionals are provided with knowledge on how best to implement SDM in complex discussion around risk, or with people living with SMI may help to increase professionals’ motivation and confidence in practice.

In our previously published studies (25, 29), most service user participants reported having not been involved in a discussion about risk or safety, which contrasts with the accounts from mental health professionals interviewed in the current study. Professionals may not have clearly highlighted the purpose of risk discussions or may have adjusted their language in ways that do not convey the nature of the conversation. Best practice guidelines (17) state that risk assessment and management should be carried out in an atmosphere of openness and transparency, as failure to communicate effectively could hinder meaningful involvement in the process. Recent evidence further supports the importance of relational approaches in the assessment and management of self-harm and suicide risk, demonstrating that therapeutic engagement and meaningful dialogue are critical components of effective practice—particularly in inpatient and emergency care settings (22).

Professionals acknowledged communication skills as vital in facilitating discussions about risk with service users, and explained that they sometimes needed to adapt the language they used i.e., discussing safety concerns rather than risk. The language of risk has been identified as a barrier to communication in other studies (54, 55). Clancy, Happell and Moxham (56) found that the language of risk was not familiar to service users and carers and suggested that a reframing of risk was necessary to reflect service users’ and carers’ experiences and understanding. This is further supported by a large cross-national study of recovery-focused mental health care planning and co-ordination where in response to feedback from their patient and public involvement group, the authors reframed and asked participants about safety rather than risk (57). Service user-led research has also found differences in perception and the language of risk used by professionals and service users. Faulkner (58), for example, found that service users reported the risk of losing their independence as a greater concern than the potential dangers or harms that are more often a concern for professionals.

Most professionals said that they were confident in discussing risk with service users. While some said their confidence was increased by training, clinical experience, team structure and MDT support.

Others believed it was limited by environmental factors such as time constraints and workload. These findings are consistent with the literature, which indicates that bureaucratic demands impede on professionals’ opportunity to meaningfully engage with service users in RA and RM (49, 53, 59, 60).

Nonetheless, professionals interviewed in this study expressed positive attitudes towards collaborating with service users in the assessment and management of risk. Their optimism derived from them wanting to support service users’ choices in the decision-making process, promoting empowerment, and providing the service user with knowledge and understanding about their risks so they were better equipped in managing them. Professionals emphasised the need to move away from the ideology that they are the’experts’ who are solely responsible for treatment, instead highlighting the role of the service user in taking ownership and responsibility for identifying and managing risk. Indeed, the model in which professionals are encouraged to share responsibility for risk is recommended in policy and research (17, 59)

### Strengths and limitations

This study identified a comprehensive list of barriers and enablers to shared decision-making in risk assessment and management. The Theoretical Domains Framework (TDF) guided both the interview schedule and data analysis, with an expert validating the belief statements generated from the study’s findings. Rigorous data analysis methods, including double coding all transcripts and inter-rater reliability checks, were employed. Another strength of this study is the high response rate (75%) and the purposive sampling strategy, which selected a diverse sample representing various disciplines, roles, and settings. This diversity enhances the transferability of the findings across different professional perspectives and service contexts.

However, the study has some limitations. First, implementing SDM in risk assessment and management encompasses a range of behaviours, each of which may encounter distinct barriers and enablers. While this study sought insights from mental health professionals about SDM as a whole, a deeper examination would require unpacking each component (i.e., informed, involved and influential) individually, which would have required significantly more time and effort from participants. Although the interviews were conducted in 2017, findings still remain relevant as the 2007 Department of Health policy on best practices in the assessment and management of risk is still in effect (17). However, recent guidelines (2, 20, 21) reflect a growing emphasis on relational approaches, particularly in the context of suicide and self-harm. This shift is supported by emerging evidence (22), reinforcing the continued relevance of the study’s themes. Third, the study primarily focused on care coordinators and senior professionals, including managers and psychiatrists.

However, the relatively small sample size, particularly within certain professional groups i.e., Psychiatrist and Occupational Therapists, may limit insight into the views and experiences specific to those roles. The service users interviewed in our previously published paper (29) emphasised the involvement of both psychiatrists and psychologists in the assessment and management of their risk, therefore further investigation is warranted to capture their specific experiences of implementing SDM with individuals with SMI.

### Implications for practice and research

To support the implementation of shared decision making (SDM) in the assessment and management of risk, enhanced training programs co-produced with service users could offer practical strategies for embedding SDM into clinical practice. Integrating this training into higher education curricula could help mental health professionals receive guidance early in their careers, addressing common anxieties around discussing risk, such as suicidality and risk to others, with service users (28, 51). However, SDM also depends on the availability of sufficient time to facilitate meaningful collaboration between service users and professionals. A cost-benefit analysis may be needed to determine the benefits of additional resources required to enable SDM. This could help inform service planning by identifying where additional capacity or structural changes are most needed to support relational, recovery-oriented approaches to risk.

There may be situations where discussing risks with service users is not possible, such as when they lack capacity, or where they are unwilling to engage in the process. However, efforts can always be made to ensure that service users are provided with the opportunity to participate in risk discussions wherever feasible. This includes informing them about the process, and offering them the chance to be involved if and when they are willing or able to do so. To support SDM, practical tools such as advance directives and crisis planning can be employed (16, 44), particularly to facilitate decision-making before a person loses capacity. Furthermore, the involvement of multidisciplinary teams and the support of other professionals are essential for the successful implementation of SDM in risk assessment and management practices.

The findings of this study may be helpful for future intervention design to enhance SDM in the assessment and management of risk, but we need to first understand interventions that have been tested to enhance service user involvement, as well as the benefits of shared decision making in these processes.

## Conclusion

The findings of the study highlight that mental health professionals support the use of SDM in the assessment and management of risk, but a range of complex factors prevent explicit and consistent implementation in practice. A key benefit of shared decision-making is its potential to enhance awareness of risks for both service users and professionals. Often, one party may be unaware of the risks that concern the other, leading to gaps in communication and a lack of openly discussed plans for effective risk management. By fostering collaborative discussions, shared decision-making ensures that both perspectives are acknowledged and addressed, enabling the development of comprehensive and mutually agreed-upon strategies for managing and minimising risks.

## Declarations

### Ethics approval and consent to participate

All procedures performed in studies involving human participants were in accordance with the ethical standards of the institutional and/or national research committee and with the 1964 Helsinki declaration and its later amendments or comparable ethical. Ethical approval was granted by the NHS Health Research Authority Research Ethics Committee London—Camden & Kings Cross (16/LO/1918), and all participants provided written informed consent.

### Consent for publication

Not applicable.

### Availability of data and materials

The data that support the findings of this study are available on request from the corresponding author. The data are not publicly available due to privacy or ethical restrictions.

### Clinical trial number

Not applicable

## Funding

This work was supported by the Wellcome Trust [315841/Z/24/Z]. Research reported in this paper was conducted as part of a PhD that was jointly funded by City, University of London, and East London NHS Foundation Trust (awarded to NA). The funders had no role in study design, data collection and analysis, decision to publish, or preparation of the manuscript.

### Authors’ contributions

All authors have read and approved the final manuscript. NA created the protocol for the study with guidance and contribution from her supervisory team AS, SB and LR. NA conducted the recruitment for the study, and interviews. NA and FB jointly coded data, and AS, SB, LR and KM contributed to the formal analysis and validation. All authors supported drafting and development of the manuscript.

## Data Availability

All data produced in the present study are available upon reasonable request to the authors

## Acknowledgements

The authors thank all the mental health professionals who took part in an interview, as well as the administrators and managers for their invaluable support throughout the project.

