## Additional file 1 for "Perceived Barriers and Enablers to Shared Decision-Making in Assessment and Management of Risk: A Qualitative Interview Study with Mental Health Professionals": Additional file 1.pdf

**Hi. My name is XXXX.**

**Thank you for kindly agreeing to take part in this interview. I am here today to ask you a few questions about risk assessments and the risk management plans. It should take about an hour. There are no right or wrong answers, we just would like to know about how you assess and manage risk for service users with severe mental health problem.**

**Confidentiality**

- Remind the person that they have already given consent to be interviewed and check they are still ok with that.
- They may stop at any time.
- Audio-recording interview.
- Remind them that their name will not be used and they will not be identifiable in any way.
- Check Dictaphone is working.
- Read out code name so that their name can be left out of the interview.

**I would like to start by asking you a few general questions about risk assessments and risk management.**

1. What does the term risk means to you?

   *Some people may focus on one aspect of risk but it can mean lots of different things. Risk can mean the possibility of any harm to the service user themselves and/or others; this may include violence, aggression, self-harm, suicide, neglect or relapse. Risk can also mean experiencing side effects from medication; harassment; stigma; discrimination, vulnerability and even isolation.*

2. Please walk me through the process of how risk would be assessed for someone who is newly accepted for treatment within your service? Would this process be any different for an **existing** service user?
3. How is risk managed? What is the risk management plan? Who develops it?
4. How often is risk assessed or managed?
5. What is your current role in relation to assessing and managing risk?
6. Are there any challenges associated to assessing and/or managing risk for individuals with severe mental health problems?

**Thank you.**

The rest of the interview will focus on more specific questions in relation to your experience of shared decision making in risk assessments and risk management planning.

7. What does shared decision making mean to you?

**Thank you**

*Shared decision making is about working in partnership with service users and/or their carer (if appropriate) and providing them with the opportunity to be <u>involved, informed</u> and <u>influential</u> in a decision making process. We are interested in how shared decision making is implemented in the process of identifying and managing risk.*

Please bear with me if some of the following questions seem repetitive, as we have used several theories about human behaviour and we would like to figure out the theory that best applies to our study.

So lets' start……..

**General Question**

8. What are your thoughts about implementing shared decision making in the assessment and management of risk?

**Knowledge**

9. Are you aware of any mental health guidelines or policies that recommend for shared decision making to be implemented in risk assessment and/or risk management planning? Yes, what is your understanding of this recommendation?

**Skills**

10. Do you think implementing shared decision making in risk assessment and/or risk management practices requires certain skills and expertise? Yes, what skills? What experience?
    *Prompts: Do you think you have these skills? What additional skills or training might you need?*

**Social /professional role and identity**

11. Whose role is it to implement shared decision making in risk assessment and risk management? Would you consider it part of your role? Why is that?
    *Prompt: would your colleagues agree?*

**Beliefs about capabilities**

12. How easy is it for you to implement shared decision in risk assessments and risk management for individuals with severe mental health problems?
13. Are there any difficulties? What would help you overcome these difficulties?
14. How confident are you about implementing shared decision making, despite these problems?

**Optimism**

15. How optimistic or pessimistic are you that in the future you will be able to implement shared decision making in risk assessments or risk management?

**Beliefs about consequences**

16. What are the benefits of implementing shared decision making in risk assessments and risk management planning? What about any potential harms or disadvantages? *Prompt: When you think about the potential benefits versus potential harms, does one outweigh the other? In what way?*

**Reinforcement**

17. Is there anything that encourages you to implement shared decision making in risk assessments and risk management practices?
    Prompt: Are there any incentives? Is there anything that discourages you?

**Intention**

18. Do you think you will implement shared decision making in future risk assessments and risk management plans?

**Goal**

19. Taking the risk assessment and risk management process as a whole, how important is it to include service users in shared decision making? Why?
    *Prompt: is it something you feel you need to do?*

**Memory, Attention and Decision Process**

20. What guides your decision of whether or not to include a service user in the process of assessing and managing their risks?
    *Prompt: what goes through your mind? What rules of thumbs do you use to reach your decision?*

**Environmental Context and Resource**

21. What aspects of your clinical environment influence whether you are able to implement shared decision making in risk assessments and/or risk management?
    *Prompts: physical vs resource factors*

**Social influence**

22. How might the views or opinions of others influence whether or not you implement shared decision making in risk assessments and/or risk management?
    *Prompt: colleagues, managers, service users, carers*

**Emotions**

23. Does shared decision making in risk assessments evoke any emotional response in you? If so, what?

**Behavioural Regulation**

24. If you wanted to increase the use of shared decision making in your risk assessments and risk management practices, how would you go about doing this? Would you make any plans? Are there any procedures you would follow?
25. Can you suggest any interventions that may help?

**END**

That's all the questions I have for you. Is there anything else you would like to say that we have not covered?

Thank you very much for your time.
