## Additional file 2 for "Perceived Barriers and Enablers to Shared Decision-Making in Assessment and Management of Risk: A Qualitative Interview Study with Mental Health Professionals": Additional file 2.docx

Additional file 2. All belief statements by domain (n=75) and the frequency with which they were coded from mental health professional interviews

| Domain | Specific belief | No. of participants | Total no. of quotes |
| --- | --- | --- | --- |
| Knowledge | I know the guidelines and/or policies that recommend SDM is implemented in RA and RM | 8 | 10 |
|  | I do not know the guidelines and/or policies that recommend SDM is implemented in RA and RM | 7 | 7 |
|  | We need to update our own knowledge and learn new ways of implementing SDM in RA and RM | 5 | 7 |
| Skills | My clinical experience has provided me with the skills I need to implement SDM in RA and RM | 11 | 16 |
|  | Implementing SDM in RA and RM with individuals with SMI is difficult/sensitive | 10 | 22 |
|  | I need more training in how best to implement SDM in RA and RM with individuals with SMI | 10 | 11 |
|  | Implementing SDM in RA and RM requires good communication skills | 8 | 12 |
|  | My professional training has prepared me for implementing SDM in risk assessment and risk management | 7 | 13 |
|  | There needs to be better communication between service users and professionals about risk | 7 | 11 |
|  | I adapt the language I use to communicate risk when implementing SDM in RA and RM | 7 | 11 |
|  | We need to improve information sharing between professionals about their experiences of implementing shared decision making in risk assessment and risk management | 7 | 9 |
|  | Therapeutic skills are important when implementing SDM in RA and RM | 4 | 5 |
|  | Implementing SDM in RA and RM requires patience | 3 | 4 |
|  | Good listening skills are important when implementing SDM in RA and RM | 3 | 3 |
|  | Good decision-making skills are important when implementing SDM in RA and RM | 2 | 4 |
|  | Empathy is important when implementing SDM in RA and RM | 2 | 2 |
|  | Emotional awareness is important when implementing SDM in RA and RM | 1 | 1 |
|  | Honesty is important when implementing SDM in RA and RM | 1 | 1 |
|  | Compassion is important when implementing SDM in RA and RM | 1 | 1 |
|  | A non-judgemental attitude is important when implementing SDM in RA and RM | 1 | 1 |
| Social/professional role and identity | The service user and the professional are jointly responsible for implementing SDM in RA and RM | 11 | 19 |
|  | The care-coordinator (i.e. the lead professional) is responsible for implementing SDM in RA and RM | 10 | 14 |
|  | It is a team responsibility to implement SDM in RA and RM | 8 | 9 |
|  | Acting in the service user's best interest can stop me from implementing SDM in RA and RM | 7 | 13 |
|  | I (we) already routinely implement SDM in RA and RM | 6 | 13 |
|  | The responsible clinician or manager is responsible for implementing SDM in RA and RM | 3 | 6 |
| Beliefs about capabilities | I feel confident about implementing SDM in RA and RM | 11 | 18 |
|  | I do not feel confident about implementing SDM in RA and RM | 4 | 7 |
| Optimism | I am optimistic that I will be able to implement SDM in RA and RM | 11 | 19 |
|  | I do not know how I feel about implementing SDM in RA and RM | 4 | 5 |
| Beliefs about consequences | I worry that talking about risk with individuals with severe mental illness may cause them distress, alarm or relapse | 9 | 21 |
|  | Implementing SDM in RA and RM with individuals with SMI can reduce or minimise risk | 9 | 13 |
|  | I worry that talking about risk with individuals with severe mental illness may cause them to disengage from services | 9 | 9 |
|  | I worry that talking about risk with individuals with severe mental illness may cause them to feel stigmatised or labelled | 7 | 19 |
|  | Implementing SDM in RA and RM promotes the service users safety | 6 | 12 |
| Reinforcement | Promoting recovery, empowerment and person-centred care encourages me to implement SDM in RA and RM | 14 | 52 |
|  | Being open and honest with service users encourages me to implement SDM in RA and RM | 13 | 31 |
|  | Safeguarding the individual and/or others from risk encourages me to implement SDM in RA and RM | 7 | 7 |
|  | Sharing responsibility with others encourages me implement SDM in RA and RM | 5 | 7 |
|  | Empathy motivates me to implement SDM in RA and RM | 3 | 3 |
|  | I feel it is important for the service user to be involved in the RA and RM process, as they are the expert in their own life | 2 | 4 |
|  | Positive risk taking encourages me to implement SDM in RA and RM | 2 | 3 |
| Intention | I will implement SDM in future RA and RM | 10 | 20 |
|  | We are working towards implementing SDM in RA and RM more regularly with service users | 2 | 4 |
| Goals | It is important to implement SDM in RA and RM | 15 | 51 |
|  | It is important to provide the service user with knowledge and understanding about their risks | 10 | 15 |
|  | It is important to discuss risk with service users for information gathering purposes | 7 | 10 |
|  | Reaching a shared decision about risk with the service user is important | 6 | 13 |
|  | Implementing SDM in RA and RM with individuals with SMI improves engagement or the therapeutic relationship | 6 | 7 |
|  | It is important to implement SDM in RA and RM to develop advance directives | 4 | 5 |
| Memory, attention and decision processes | The type/level of risk influences my decision to implement SDM in RA and RM | 11 | 29 |
|  | Implementing SDM in RA and RM with an individual with SMI is dependent on the situation | 9 | 43 |
| Environmental context and resources | The RA or RM process enables me to implement SDM in RA and RM with individuals with SMI | 11 | 25 |
|  | I do not have enough time to implement SDM in RA and RM | 8 | 25 |
|  | My team structure or setting type makes it easy to implement SDM in RA and RM | 7 | 24 |
|  | The place of meeting makes it difficult to implement SDM in RA and RM | 6 | 12 |
|  | We need more resources and time to implement SDM in RA and RM | 5 | 9 |
|  | Organisational and procedural changes need to happen to improve SDM in RA and RM | 4 | 5 |
| Social influences | The service users’ capacity is a key factor in whether I can implement SDM in RA and RM | 15 | 35 |
|  | The service users’ insight, presentation or understanding are key factors in whether I can implement SDM in RA and RM | 14 | 35 |
|  | The service users’ level of engagement is a key factor in whether I can implement SDM in RA and RM | 14 | 41 |
|  | I work as part of a multi-disciplinary team that support me to implement SDM in RA and RM | 13 | 34 |
|  | If the service user and I disagree about the risk, it can be difficult to implement SDM in RA and RM | 12 | 22 |
|  | Family members and carers help me implement SDM in RA and RM | 8 | 15 |
|  | Implementing SDM in RA and RM depends on the quality of the therapeutic relationship | 8 | 16 |
|  | Family members and carers make it harder to implement SDM in RA and RM | 7 | 9 |
|  | Interprofessional conflicts and power dynamic can make it harder to implement SDM in RA and RM | 5 | 9 |
|  | A change in staff culture is needed around implementing SDM in RA and RM with individuals with SMI | 5 | 8 |
|  | Language barriers can make it difficult to implement SDM in RA and RM | 2 | 5 |
| Emotions | Implementing SDM in RA and RM with individuals with SMI makes me feel anxious | 7 | 10 |
|  | I fear for my personal safety when implementing SDM in RA and RM | 6 | 15 |
|  | Discussing risk with service users can be uncomfortable | 4 | 7 |
|  | Manging my own emotions helps me implement SDM in RA and RM | 3 | 3 |
|  | Implementing SDM in RA and RM makes me feel positive | 3 | 3 |
| Behavioural regulation | An App to help engage service users in RA and RM | 1 | 2 |
