## Supplementary material for "Perceived Barriers and Enablers to Shared Decision-Making in Assessment and Management of Risk: A Qualitative Interview Study with Mental Health Professionals": Table 1: Table 1. Belief statements and sample quotes for each domain.docx

Table 1. Definitions of the 14 TDF domains (Cane et al, 2012), belief statements and illustrative quotes

| Domain/definition | Specific belief | Sample quote | No. of participants | Total no. of quotes |
| --- | --- | --- | --- | --- |
| Knowledge  *An awareness of the existence of something* | I know the guidelines and/or policies that recommend SDM is implemented in RA and RM | *“Yes, the Care Act 2014 yes, we need to consider service users and their carers in our decision making, risk assessment”* | 8 | 10 |
|  | I do not know the guidelines and/or policies that recommend SDM is implemented in RA and RM | *“No but there probably is some out there, I would imagine there’s quite a lot out there”* | 7 | 7 |
| Skills  *An ability or proficiency acquired through practice* | My clinical experience has provided me with the skills I need to implement SDM in RA and RM | *“I think risk assessment comes with experience and you can teach it to people but sometimes they have to live that scenario in terms of going to an assessment and until you’re qualified and in the job, you can sit in a college and learn about it but until you can relate theory to practice…”*  *“I think it requires experience. I think it also requires somebody to be quite knowledgeable and I think it requires someone that is prepared to include, be inclusive because some, I’ve worked with some people who may have some old fashioned views and it’s about challenging that and ensuring that all the staff are on board, not here but I’m going back in time now, but certainly in the last few years people have become very knowledgeable about service user involvement, carer involvement and ensuring that that’s at the centre of everything we do”* | 11 | 16 |
|  | Implementing SDM in RA and RM with individuals with SMI is difficult/sensitive | *“…in terms of people that are under our service, obviously, we try and discuss the risk regularly with them and look at how we can either help them to manage that risk. Some people can engage in those sort of conversations. Other people are like, no, this is my past, why are you bringing it up? What's this got to do with now, that was five years ago? And I think it's sometimes difficult having that conversation to explain to them, well, yes, this is your past, but also there might be elements of that we still need to be aware of...* | 10 | 22 |
|  | I need more training in how best to implement SDM in RA and RM with individuals with SMI | *“Training? I suppose building on what I already have is really important to me”*  *“I think the most powerful training is when you do it together at a team and you’re doing it with the people that work around you, because that’s when you can all relate to each other and relate to the client group that you’re working with”* | 10 | 11 |
|  | Implementing SDM in RA and RM requires good communication skills | *“Well I think being able to communicate is important, but also being able to listen, the two things are obviously related, but I think there’s also something about being able to judge the level that you provide the information at”* | 8 | 12 |
|  | My professional training has prepared me for implementing SDM in risk assessment and risk management | *“…we’re supposed to have those skills as Care Coordinators and we've done the… the risk management, assessment courses etc”*  *“… I think that my training as an AMHP really helped because a lot of that focus is around assessing risk and how you weigh it up…”* | 7 | 13 |
|  | We need to improve information sharing between professionals about their experiences of implementing shared decision making in risk assessment and risk management | *“Speak about it in team meetings, speak about it when you’re with colleagues, share experiences with colleagues, talk about advantages and disadvantages of it”* | 7 | 9 |
|  | There needs to be better communication between service users and professionals about risk | *“it would be helpful perhaps to be a bit more explicit, so using the word risk with patients, maybe thinking in the CPA… we could have a question so I could say to the patient, do you think that there are any risks?...”* | 7 | 11 |
|  | I adapt the language I use to communicate risk when implementing SDM in RA and RM | *“…we want to try and ascertain what are the risks. So like you said earlier, it's thinking about risk to self, risk to others, vulnerabilities, risks from others, risks to children or any other vulnerable adults, and breaking it down into those four sort of categories and basically trying to ask questions. Sometimes it’s appropriate to ask those questions directly, and be quite matter of fact about it; other times you want to ease it into a conversation and not be so direct with it, and maybe sort of tease those answers out a bit.”* | 7 | 11 |
| Social/professional role and identity  *A coherent set of behaviours and displayed personal qualities of an individual in a social or work setting* | The service user and the professional are jointly responsible for implementing SDM in RA and RM | *“Everybody’s, including the patient. I do think that they also have a role in wanting to be involved and some people don’t, some people will sit there and say, I don’t want to be involved in this, I want you to do it and some people won’t say that but they will behave in a way that implies that. But I think that we should be moving in our society towards the idea that doctors and nurses aren’t responsible for treating, aren’t just responsible for treating patients, patients are also responsible for engaging in that process, it’s not all one way”*  *“Because it’s pointless us coming in and saying right we’ve identified these as the risk, we’re addressing that with you, you’re either agreeing or you’re not agreeing but we think you should do X, Y and Z and the service user going yeah and then they don’t do anything about it. So we all have to take responsibility for the way we are, the way we act, the way we respond to people and for our actions and if someone’s not taking ownership of that you could just talk till you’re blue in the face, so I think it’s a joint responsibility”* | 11 | 19 |
|  | The care-coordinator (i.e. the lead professional) is responsible for implementing SDM in RA and RM | *“It’s the role of the Care Coordinator or the professional that is the lead professional involved in that person’s life”* | 10 | 14 |
|  | It is a team responsibility to implement SDM in RA and RM | *“Everybody who works with the patient. It’s their role”*  *“I guess the person who knows the service user best, like a care co-ordinator, they can invite them along and the family, and then just the wider MDT need to facilitate it”* | 8 | 9 |
|  | Acting in the service user's best interest can stop me from implementing SDM in RA and RM | *“…But ultimately there’s a point where obviously if I was so concerned I would end up probably having to take action on their behalf, but then that’s a challenge as well in terms of when I step in as the responsible clinician to actually take responsibility for that risk, I take it away from the person…”*  *“I think it just depends on the individual and at what stage they are in their illness because some people may be getting over the worst of it if you like and then more able to have these conversations whereas others are in the thick of it and can’t really think beyond what’s going on right now so you would do what you feel is in their best interest.  Obviously with guidance and support from others that know them, like their family”* | 7 | 13 |
|  | I (we) already routinely implement SDM in RA and RM | *“Well I actually think it’s quite easy here because there’s a team of people here who are very on board with shared working practices and are very clear that service users should be included in the work that they do”*  *“we all think about this every day, and it's very much a part of our practice.”* | 6 | 13 |
| Beliefs about capabilities  *Acceptance of the truth, reality, or validity about an ability, talent, or facility that a person can put to constructive use* | I feel confident about implementing SDM in RA and RM | *“… I would like to think and I’m fairly confident that if I’ve got a service user with fluctuating risks and very significant risks then that is certainly something that I would address”* | 11 | 18 |
|  | I do not feel confident about implementing SDM in RA and RM | *“I feel less confident at talking about some of the potentially more judgey bits of risk, so them perhaps doing harm to others and I think one of the reasons for that is because I don’t want to instil the idea that because somebody’s got a diagnosis of a severe and enduring mental illness they then become a risk to others…”* | 4 | 7 |
| Optimism  *The confidence that things will happen for the best or that desired goals will be attained* | I am optimistic that I will be able to implement SDM in RA and RM | *“Fairly positive, because it's happening”*  *“I think it makes feel quite happy that they’re taking things on board and that you’re more hopeful because it’s about taking responsibility and having a plan in place”* | 12 | 21 |
| Beliefs about consequences  *Acceptance of the truth, reality, or validity about outcomes of a behaviour in a given situation* | I worry that talking about risk with individuals with severe mental illness may cause them distress, alarm or relapse | *“Upsetting them. Setting them back or encountering resistance because they don't want to talk about that particular, those particular aspects or that particular time of their life”* | 9 | 21 |
|  | Implementing SDM in RA and RM with individuals with SMI can reduce or minimise risk | *““…without meaningfully engaging with the client then the opportunity to manage risk is greatly reduced and sometimes it is just committed to a document rather than a process of interaction with the service user…”* | 9 | 13 |
|  | I worry that talking about risk with individuals with severe mental illness may cause them to disengage from services | *“…I think it’s also difficult because at the end day if you’ve got a good relationship do you really want to damage it by bringing up really painful and obviously very difficult topics with them, so it’s around timing, as well, how prepared they are to talk about it…”*  *“…people not wanting to engage with services because they’ve found out that you know, you might know something about their risk history or you know about criminal convictions and they don’t, maybe they don’t want their parents finding out”*  *“And then the other main problem I have I guess is disengagement, so potentially offending somebody or doing something in a way that means that the patient doesn’t feel that we can engage them”* | 9 | 9 |
|  | I worry that talking about risk with individuals with severe mental illness may cause them to feel stigmatised or labelled | *“There are challenges, because of the way in which risk is viewed, really. So, risk being equivalent to an understanding or a profile of dangerousness, for example. And that can be very stigmatising for the person, for the individual involved. And I always try to be aware of issues around the person’s race, around their gender, around the actual diagnosis, around other organisations that may be involved or other services that may be involved. Because, as much as we, it might be helpful for us to paint, get a profile of somebody’s risk, just to focus on that risk can be quite, I think it can be quite limiting for that person”*  *“I think it’s very difficult, particularly with risk assessment because I think that a lot of the time a lot of the things that we think are important about risk assessment are often bad things that have happened to people in the past and we make judgments about what might happen in the future on the basis of that, and people don’t like talking about those things obviously often and it’s, so it’s difficult to go through those things. If people have committed assaults when they’ve been ill or just in a different phase in their life and now they’ve turned things around, to keep going back to it as the basis for decision making in the future it’s really hard”* | 7 | 19 |
|  | Implementing SDM in RA and RM promotes the service users safety | *“… you can have a frank discussion about safety and they might feel more safe if they're aware and can make their own risk management plan with the staff. And carers might feel more supported by that. And it might help, as well, with making a relapse prevention plan, so that could be a benefit”* | 6 | 12 |
| Reinforcement  *Increasing the probability of a response by arranging a dependent relationship, or contingency, between the response and a given stimulus* | Promoting recovery, empowerment and person-centred care encourages me to implement SDM in RA and RM | *“…It’s about them. I can’t make decisions about them and they’re not included in it. It’s just going to be a statement on a piece of paper. If they agree with it they’re more likely to work with you instead against you”*  *“…I think as we’re moving towards more recovery we’re focusing more on recovery and it should be part of their recovery plan, in terms, to get them on board and feel part of it, and to take for them, service users kind of take ownership, otherwise it’s something that we’re doing to them and it’s another piece of paper that we hand to them or subject them to another assessment. And when they go through the hospital process, the admission, the discharge, they’ve had so many assessments this is, they must be fed up. And for them, you have to involve them because otherwise it’s got no meaning to them, it’s just another piece of paper or another discussion that’s just discarded”*  *“Well I think that is part of the model of early intervention. Recovery focus is a big thing at the moment and I’m supportive of that, but without wanting to blow our own trumpets we’ve been, we’ve had a recovery focus for ten years because we’re early intervention and that’s what, that’s how the model was set up in the first, before my time, I can’t take credit for that, but I think that has always been our focus, that if you can empower people to take responsibility for managing their own mental health you end up with better outcomes”* | 14 | 52 |
|  | Being open and honest with service users encourages me to implement SDM in RA and RM | *“Well it’s about them isn’t it, it’s their risk or risk that might be presented to them so they have to be as much as possible at the heart of the decision making and being open and transparent with them about that where it’s appropriate”*  *“You need to have an open dialogue with people and be honest. I think honesty is the key and definitely something that my service users that I’ve worked with, will say, is, just tell me how it is and I think that’s so important because sometimes they need to hear it as it is, no fluffiness, no making it sound nice because it’s not nice, it’s not, it is what it is and we need to try and find a way forward”* | 13 | 31 |
|  | Safeguarding the individual and/or others from risk encourages me to implement SDM in RA and RM | *“Just so that they're aware of the risk. I guess that they know that we're trying to protect their safety, that they feel supported, that their carers feel supported”*  *“Or maybe they’re walking around with knives and stuff because we do have clients that walk around with knives because they’re paranoid. You’re making them aware that look you’re informing them and saying look you’re walking around with a knife you could get stopped by the police, you could accidentally harm someone else even though you have no intention of doing that, you need to stop doing that…”* | 7 | 7 |
| Intention  *A conscious decision to perform a behaviour or a resolve to act in a certain way* | I will implement SDM in future RA and RM | *“Usually I discuss the risk with every service user, yes”*  *“Yeah I think, yeah, we wouldn’t want to go the other way, so I think we’d like to, yeah, to try and keep doing that”* | 10 | 20 |
| Goals  *Mental representations of outcomes or end states that an individual wants to achieve* | It is important to implement SDM in RA and RM | *“… you have to involve the service user regardless because you know otherwise it makes it just a paper exercise and it becomes meaningless”*  *“…But I do think it’s vital to involve them, because if we can involve them and allow them to have a say in what the future care might be, then, again, they also need to be, able to be or have a say in what the risk plans are and the risk management plans might be”* | 15 | 51 |
|  | It is important to provide the service user with knowledge and understanding about their risks | *“They have information. They know that if they continue to allow certain people into their homes they can lose their flat. They know that sometimes when people take drugs they behave in chaotic ways and their lifestyles get chaotic and there could be violence towards them from others. They understand because they’ve been down that very long road. They know that that sometimes their mental health if they’re ill they present as quite violent because of the voices they hear and what they see”* | 10 | 15 |
|  | It is important to discuss risk with service users for information gathering purposes | *“…during that assessment of somebody we would be looking to gather information from them about their understanding of the risks that they experience, so any issues that they might have with being vulnerable or issues related to their social circumstances or their psychological or mental wellbeing that might put them at risk of coming to some harm, or the people around them coming to harm”* | 7 | 10 |
|  | Reaching a shared decision about risk with the service user is important | *“In any moment or even with the best or good intentions you may get it wrong. However, the probability of getting things wrong will be less if it’s a shared decision…”* | 6 | 13 |
|  | Implementing SDM in RA and RM with individuals with SMI improves engagement or the therapeutic relationship | *“…if they're involved in a shared risk management it’s much better. It’s much, much better than actually working, working or putting something in place whereby they don't really understand and they don't want to engage with you which can be difficult…”*  *“With regards to risk, as long as you inform them and you update them I think they’re quite happy and it makes it easier to actually work with them in the long run rather than doing things where they’re not fully cognisant with it. When they’re fully cognisant with what’s happening, whether good or bad, they’re a lot easier to work with in that sense”* | 6 | 7 |
| Memory, attention and decision processes  *The ability to retain information, focus selectively on aspects of the environment and choose between two or more alternatives* | The type/level of risk influences my decision to implement SDM in RA and RM | *“But it depends on the risk. If it’s a positive risk that the person wants to take then yes, I’m fine with it, I’m OK with it, but if it’s a negative risk as in a risk where there’s a potential, the potentiality of harming himself or harming other people is there and I need to discuss it. Then it will make me a bit wary and what I will do in that sense is I’ll get someone else to, a senior officer, senior practitioner to sit with me while I’m discussing with this person”*  *“I suppose the riskier someone is the more likely I am to include them in the process. I’ve got a couple of people who don’t seem to pose an overt risk to themselves or others and so it’s much less of a discussion”* | 11 | 29 |
|  | Implementing SDM in RA and RM with an individual with SMI is dependent on the situation | *“Not necessarily because each situation is unique in itself, and each decision you make will not be the same because everybody’s an individual. So, regardless it might have the same problem, but the outcome of each problem would differ depending on each person’s situation. So, I don’t think one outweighs the other it has to be based on each individual, on the approach you need for each individual basically”*  *“I think with all the will in the world or the procedures it’s based on each individual service user and their circumstance, so although you can say right this is how you manage violence or this is how you manage this, it’s very much individual to each person and how on board they’re happy to be and how open they’re happy to be around their risk and the management of that risk. So, I would take it case by case rather than saying right in early intervention we can do this with everybody, we can’t you’ve got to adapt it for each person…”* | 9 | 43 |
| Environmental context and resources  *Any circumstance of a person's situation or environment that discourages or encourages the development of skills and abilities, independence, social competence, and adaptive behaviour* | The RA or RM process enables me to implement SDM in RA and RM with individuals with SMI | *“Once the patient’s in red we then implement this process and then the only person that can take them out of red is the doctor that’s seeing them with the care coordinator. So, once they’re in red they have to be reviewed by a doctor, at that review the risk will be reviewed. If it turns out that actually it’s OK and it, there’s no increase in risk then they’d go back into amber. If they remain at risk then we would keep them in red for a bit longer and then they’d be reviewed again within the month. That’s the plan, it’s not failsafe”* | 11 | 25 |
|  | I do not have enough time to implement SDM in RA and RM | *“The other thing is the lack of time, lack of time essentially and getting everybody together and using that opportunity to look at those risks”*  *“If the person did engage, obviously, that would be useful. Trying to promote engagement is time consuming, so a lower caseload helps with that. I think that’s part of the capacity for more close observation and attention to risk in early intervention services because they traditionally have lower caseloads. We’re looking at about 15 to 20 rather than 20 to 30”* | 8 | 25 |
|  | My team structure or setting type makes it easy to implement SDM in RA and RM | *“the relapse prevention plans that I’ve outlined, I’ve been working in early prevention now for a number of years, but I think that they are quite specific to EI. So, most teams don’t really look in such detail at early, middle and late warning signs and they probably involve the client a bit less in joint decision making. They don’t have such regular meetings as we do, as I said we have one every morning and so for professionals I think it’s the frequency of meetings and the openness of discussions about risk in teams and the willingness for people in, across different roles to take a role and responsibility in the management of risk”* | 7 | 24 |
| Social influences  *Those interpersonal processes that can cause individuals to change their thoughts, feelings, or behaviours* | The service users’ capacity is a key factor in whether I can implement SDM in RA and RM | *“… if the person’s unwell, very unwell where they need to be hospitalised, sitting there, discussing risk is a waste of time basically for yourself and for them. They need to be treated, they need to be in hospital, they need to be in a place of safety so they’re not a risk for themselves or others so yeah”*  *“I guess the only circumstances that it might not be discussed is if they don't have capacity to understand at that time”* | 15 | 35 |
|  | The service users’ insight, presentation or understanding are key factors in whether I can implement SDM in RA and RM | *“… if someone lacks insight obviously, then that’s a barrier to assessing risk because you can’t have that joint agreement of the problem. So, you can’t come to a shared understanding of how to manage risk”* | 14 | 35 |
|  | The service users’ level of engagement is a key factor in whether I can implement SDM in RA and RM | *“sometimes it’s difficult because they don’t want to engage”*  *“It’s vital if you’re managing risk successfully, but equally if someone is uninterested or unwilling to be part of that process then there’s nothing you obviously can do to force it, really. Then it is reliant on the professionals and building documents and plans in the absence of service user involvement”* | 14 | 41 |
|  | I work as part of a multi-disciplinary team that support me to implement SDM in RA and RM | *“Having access to the right people at the right time. So not after somebody’s actually had the crisis and there’s been a gap it should be, they've had the crisis, period of recovery and then talking about those particular risks. And it should be shared so that client doesn't think that only one worker for example the Care Coordinator is fixated on, or just keeps harping on about the risk”*  *“Because we try and make our service users aware that we do have a team approach, so although they’ll have a regular person that they’re meeting with they might be meeting with other people, whether that’s through the care coordinator being on holiday or some of our activity groups, so they get to know different people within the team”* | 13 | 34 |
|  | If the service user and I disagree about the risk, it can be difficult to implement SDM in RA and RM | *“Some service users are just point, deny bluntly deny that that ever happened, so that puts you in a difficult position as well”*  *“… they don’t always take advice, they don’t always do what they say they’re going to do and so you think you’ve got a plan in place and then it can go pear shaped because some step hasn’t been taken that you can’t make people do things…”* | 12 | 22 |
|  | Family members and carers help me implement SDM in RA and RM | *“… the family also can identify the risk and how they can reduce the risk”*  *“This is about naming the risk and trying to find ways to minimise it and that’s normally done again using the service user, family, carers and any other people that may be already involved in that network to try and manage the risk”* | 8 | 15 |
|  | Family members and carers make it harder to implement SDM in RA and RM | *“he wouldn’t report them because I don’t want to get my brother in trouble. And, and it was hard to make him see well this is a safeguarding issue, this is someone taking advantage of you…”* | 7 | 9 |
|  | Implementing SDM in RA and RM depends on the quality of the therapeutic relationship | *“Yeah it does require skills, and it also required having a good relationship with the patient themselves if there isn't a trusting relationship there then it’s going to be very difficult to talk about very personal, very traumatic times and things that they’ve done in the past or the way they’ve behaved or how they presented. It’s, it can open up a lot of wounds, so you have to be prepared to kind of deal with the emotional aspect of that in terms of discussing those particular risks”* | 8 | 16 |
| Emotions  *A complex reaction pattern, involving experiential, behavioural, and physiological elements, by which the individual attempts to deal with a personally significant matter or event* | Implementing SDM in RA and RM with individuals with SMI makes me feel anxious | *“I think it can maybe make some staff feel anxious, if that’s something that they don’t have a lot of experience with, I think for me I’ve worked for a few years but sitting down and discussing risk with someone, I wouldn’t expect anyone to say yeah, no problem, I’ll do that. I think as a person and I think naturally if you don’t go into different situations around risk, feeling slightly nervous or slightly anxious that would worry me in a way”* | 7 | 10 |
|  | I fear for my personal safety when implementing SDM in RA and RM | *“…But obviously you’ve got to think about safety for yourself and if you’re going to be having a conversation about risk with a service user you need to think well where’s the most, when’s the most appropriate time to do that in terms of their mental state, what’s happening in their life at the time, where’s the most appropriate place to do that?”*  *“How confident? If someone was shouting and screaming about it I wouldn’t be very confident if I’m on my own but if I’m with another member of staff I think I would be quite confident but not on my own. I wouldn’t put myself in danger. If a person is not ready to receive the information because they’re agitated or can be violent towards you then I won’t be sharing the information with them. I’ll have to think of my safety first”* | 6 | 15 |
| Behavioural regulation  *Anything aimed at managing or changing objectively observed or measured actions* | An App may help to engage service users in RA and RM | *“…I think that will be a great thing if there was an app on a phone that, or an opportunity for the service user to truly engage in feeding into that risk assessment and having full access to it”* | 1 | 2 |
