## Supplementary material for "Perceived Barriers and Enablers to Shared Decision-Making in Assessment and Management of Risk: A Qualitative Interview Study with Mental Health Professionals": Table 2: Table 2. Mental health professional participants’ characteristics.docx

Table 2. Characteristics of Mental Health Professional Participants (n=15)

| Characteristic | Senior Practitioners (n = 4) | Care Coordinators (n = 11) |
| --- | --- | --- |
| Age^1^, mean years (SD), range | 51 (7), 43–59 | 37 (6), 28–46 |
| Gender, % (N) | Female: 75 (3)  Male: 25 (1) | Female: 82 (9)  Male: 18 (2) |
| Ethnicity, % (N) | White (UK/Irish): 50 (2)  White (Other): 25 (1)  Black African: 25 (1) | White (UK/Irish): 18 (2)  Bangladeshi: 18 (2)  Asian (Other): 18 (2)  Black African: 36 (4)  Black Caribbean: 9 (1) |
| Professional Role, % (N) | Mental Health Nurse / Psychiatrist: 100 (4)* | Mental Health Nurse: 36 (4)  Social Worker: 45 (5)  Occupational Therapist: 18 (2) |
| Highest Education Level , % (N) | Data suppressed to preserve anonymity* | Degree: 55 (6)  Masters: 36 (4)  Postgraduate Diploma/Certificate: 9 (1) |
| Years working in Mental Health, % (N) | 10+ years: 100 (4) | 10+ years: 45 (5)  7–9 years: 18 (2)  4–6 years: 18 (2)  1–3 years: 18 (2) |
| Years in Current Service, % (N) | 10+ years: 75 (3)  4–6 years: 25 (1) | 10+ years: 27 (3)  4–6 years: 18 (2)  1–3 years: 36 (4)  <1 year: 18 (2) |

*Senior Practitioner roles combined to preserve participant anonymity (Mental Health Nurse and Psychiatrist).

Missing Data: ^1^Age, senior practitioner (N = 1), care coordinators (N = 3)

All values represent % (n) or (standard deviation) and range.
